# The potential impact of the COVID-19 pandemic on the tuberculosis epidemic – a modelling analysis

**DOI:** 10.1101/2020.05.16.20104075

**Authors:** Lucia Cilloni, Han Fu, Juan F Vesga, David Dowdy, Carel Pretorius, Sevim Ahmedov, Sreenivas A.Nair, Andrei Mosneaga, Enos Masini, Suvanand Sahu, Nimalan Arinaminpathy

**Author notes:** Correspondence to*: Nimalan Arinaminpathy.

## Abstract

**Background:** Routine services for tuberculosis (TB) are being disrupted by stringent lockdowns against the novel SARS-CoV-2 virus. We sought to estimate the potential long-term epidemiological impact of such disruptions on TB burden in high-burden countries, and how this negative impact could be mitigated.

**Methods:** We adapted mathematical models of TB transmission in three high-burden countries (India, Kenya and Ukraine) to incorporate lockdown-associated disruptions in the TB care cascade. The anticipated level of disruption reflected consensus from a rapid expert consultation. We modelled the impact of these disruptions on TB incidence and mortality over the next five years, and also considered potential interventions to curtail this impact.

**Results:** Even temporary disruptions can cause long-term increases in TB incidence and mortality. We estimated that a 3-month lockdown, followed by 10 months to restore normal TB services, would cause, over the next 5 years, an additional 1.65 million TB cases (Crl 1.49– 1.85) and 438,000 TB deaths (CrI 403 – 483 thousand) in India, 41,400 (28,900–62,200) TB cases and 14,800 deaths (10.5 – 19.2 thousand) in Kenya, and 7,960 (6,250 – 9,880) cases and 2,050 deaths (1,610 – 2,360) in Ukraine. However, any such negative impacts could be averted through supplementary “catch-up” TB case detection and treatment, once restrictions are eased.

**Interpretation:** Lockdown-related disruptions can cause long-lasting increases in TB burden, but these negative effects can be mitigated with targeted interventions implemented rapidly once lockdowns are lifted.

## Introduction

The emergence of the novel virus SARS-CoV-2 has caused morbidity, mortality and societal disruption on a global scale. In the absence of pharmaceutical interventions, many countries have resorted to population-wide lockdowns to slow the spread of the virus and to allow their health systems to cope^1^. These lockdowns have had an important effect on SARS-CoV-2 transmission^2,3^. However, unintended consequences are inevitable with such sweeping measures. In low- and middle-income countries with health systems already under strain, even temporary disruptions in health services can have lasting impact on population health^4,5^.

In the present study we focus on tuberculosis (TB) – globally, the leading cause of death due to an infectious disease^6^. In recent decades TB incidence and mortality have been steadily declining, reflecting ongoing improvements in diagnosis, treatment and prevention^7^. However, in March 2020 a rapid analysis conducted by the Stop TB Partnership brought attention to severe impacts of COVID-related lockdowns on TB care in different countries^8^. For example, in the weeks following the imposition of a nationwide lockdown on March 24, 2020, India reported an 80% drop in daily notifications of TB^8^ relative to average pre-lockdown levels. Such declines, likely reflecting reductions in access to diagnosis and treatment, could have a lasting impact on TB burden at a country-wide level. Missed diagnoses would mean increased opportunities for transmission, while worsened treatment outcomes increase the risk of death from TB. Therefore, while lockdowns are an important measure to mitigate the immediate impact of COVID–19, it is critical to anticipate (i) the potential long-term impact of these measures on TB and other diseases, and (ii) how this impact might be stemmed, in the short term, by appropriately targeted investment and effort. We therefore aimed to examine these questions using mathematical modelling of TB transmission dynamics. Building on earlier modelling conducted for the 2019 Lancet Commission on Tuberculosis^9,10^, we modelled the potential TB-related impact of COVID-related lockdowns – and mitigating effects of potential post-lockdown interventions – in three focal countries: India, the Republic of Kenya, and Ukraine.

## Methods

### Model overview

For each country we drew from previously published models of TB transmission^10^, which were designed to capture essential features of the TB care cascade. For the current analysis, this approach allowed us to model the impact of disruptions acting at multiple points in the care cascade. For India we incorporated the dominant role of the private healthcare sector in providing TB care^11^; for Kenya, the role of HIV in driving TB dynamics^12^; and for Ukraine, the burden of drug resistance^13^. We calibrated each country model to the available data on TB burden, including WHO estimates of TB incidence and mortality^6^, and on the burden of drug resistance. Full details of each model are provided in the Supporting Information.

Calibration was performed using Markov Chain Monte Carlo (MCMC) simulation^14–16^, whereby we allowed model parameters to vary over pre-specified prior distributions, using a likelihood function based on the calibration targets listed above to weight simulations according to their fit to the observed data. For each country, we drew 250 samples from the weighted (posterior) density of simulations following burn-in and thinning as described in the Supporting Information. We then performed model projections on the basis of each of these samples, under the lockdown scenarios described below. For any model projection (for example, incidence over time), we estimated Bayesian credible intervals as 2.5^th^ and 97.5^th^ percentiles, and central estimates as 50^th^ percentiles, of the corresponding posterior density.

### Modelling the impact of disruptions

Disruptions to TB services can act at all stages of the TB care cascade. During a lockdown, movement restrictions would curtail opportunities for those experiencing TB symptoms to seek care. Even once these people are able to visit a provider or health facility, the diagnostic and laboratory capacity needed to support TB diagnosis may be severely reduced – for example, with molecular diagnostic tools for TB being repurposed for COVID–19e^17^ or TB laboratory staff being redirected to Covid-19 efforts. National TB programmes are investing significant effort to continue supporting those already on TB treatment, but there are also concerns that lockdown conditions may interfere with the continued supply of drugs^18^. To capture this range of possible disruptions, we performed a rapid consultation amongst experts at the Stop TB Partnership and the United States Agency for International Development (USAID). Table 1 lists those experts’ consensus opinion as to the degree to which TB services could be disrupted by COVID-related lockdowns, at each step of the care cascade. There is substantial uncertainty around these possible impacts, and as described below, we performed sensitivity analysis to identify which components of disruptions would have the greatest impact on overall TB burden.

**Table 1.**
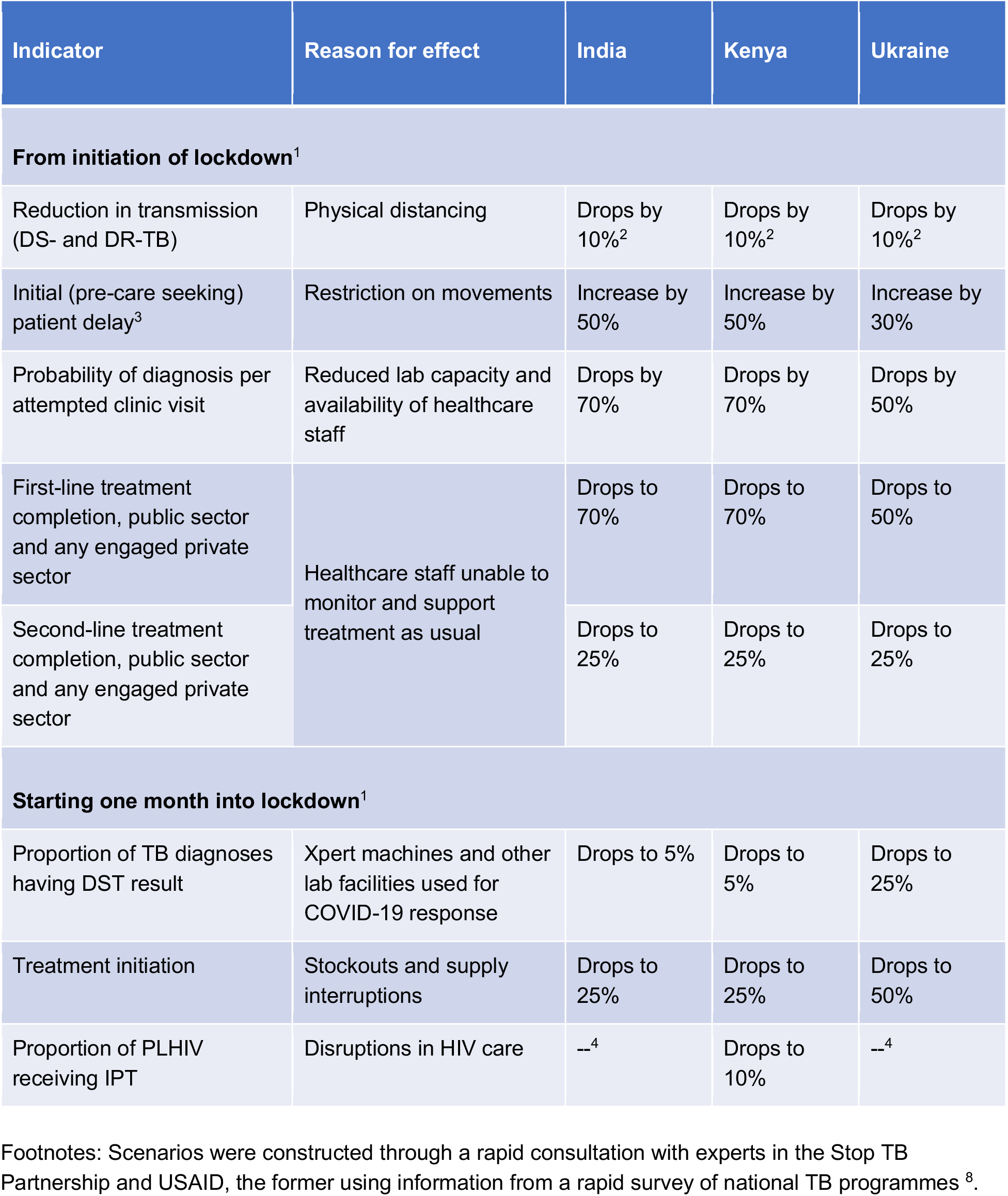
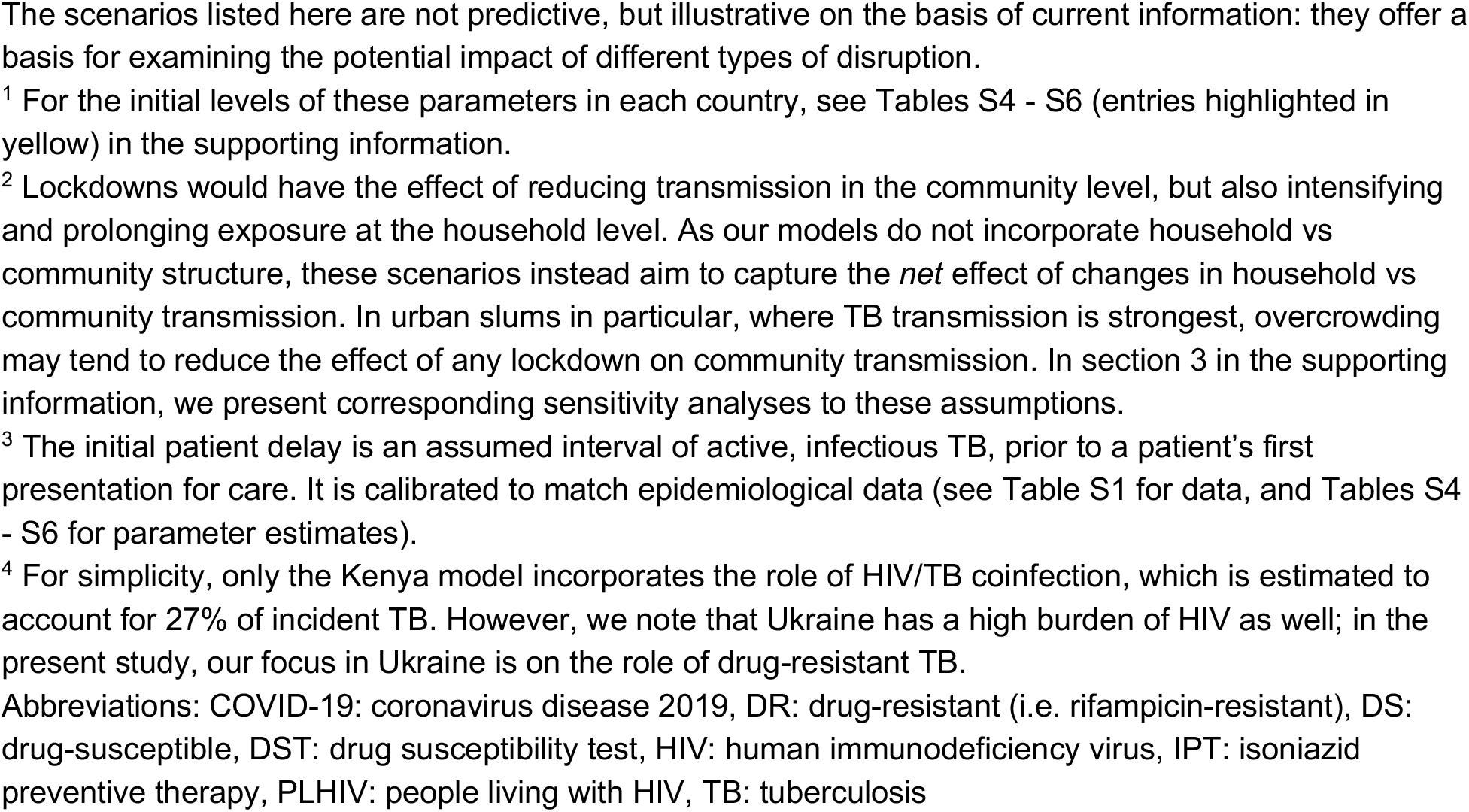
Expert consensus on potential disruptions arising from COVID-related lockdowns in three countries.

| Indicator | Reason for effect | India | Kenya | Ukraine |
| --- | --- | --- | --- | --- |
| From initiation of lockdown<sup>1</sup> |  |  |  |  |
| Reduction in transmission (DS- and DR-TB) | Physical distancing | Drops by 10%<sup>2</sup> | Drops by 10%<sup>2</sup> | Drops by 10%<sup>2</sup> |
| Initial (pre-care seeking) patient delay<sup>3</sup> | Restriction on movements | Increase by 50% | Increase by 50% | Increase by 30% |
| Probability of diagnosis per attempted clinic visit | Reduced lab capacity and availability of healthcare staff | Drops by 70% | Drops by 70% | Drops by 50% |
| First-line treatment completion, public sector and any engaged private sector | Healthcare staff unable to monitor and support treatment as usual | Drops to 70% | Drops to 70% | Drops to 50% |
| Second-line treatment completion, public sector and any engaged private sector |  | Drops to 25% | Drops to 25% | Drops to 25% |
| Starting one month into lockdown<sup>1</sup> |  |  |  |  |
| Proportion of TB diagnoses having DST result | Xpert machines and other lab facilities used for COVID-19 response | Drops to 5% | Drops to 5% | Drops to 25% |
| Treatment initiation | Stockouts and supply interruptions | Drops to 25% | Drops to 25% | Drops to 50% |
| Proportion of PLHIV receiving IPT | Disruptions in HIV care | --<sup>4</sup> | Drops to 10% | --<sup>4</sup> |

Footnotes: Scenarios were constructed through a rapid consultation with experts in the Stop TB Partnership and USAID, the former using information from a rapid survey of national TB programmes <sup>8</sup>.
The scenarios listed here are not predictive, but illustrative on the basis of current information: they offer a basis for examining the potential impact of different types of disruption.
<sup>1</sup> For the initial levels of these parameters in each country, see Tables S4 - S6 (entries highlighted in yellow) in the supporting information.
<sup>2</sup> Lockdowns would have the effect of reducing transmission in the community level, but also intensifying and prolonging exposure at the household level. As our models do not incorporate household vs community structure, these scenarios instead aim to capture the *net* effect of changes in household vs community transmission. In urban slums in particular, where TB transmission is strongest, overcrowding may tend to reduce the effect of any lockdown on community transmission. In section 3 in the supporting information, we present corresponding sensitivity analyses to these assumptions.
<sup>3</sup> The initial patient delay is an assumed interval of active, infectious TB, prior to a patient's first presentation for care. It is calibrated to match epidemiological data (see Table S1 for data, and Tables S4 - S6 for parameter estimates).
<sup>4</sup> For simplicity, only the Kenya model incorporates the role of HIV/TB coinfection, which is estimated to account for 27% of incident TB. However, we note that Ukraine has a high burden of HIV as well; in the present study, our focus in Ukraine is on the role of drug-resistant TB.
Abbreviations: COVID-19: coronavirus disease 2019, DR: drug-resistant (i.e. rifampicin-resistant), DS: drug-susceptible, DST: drug susceptibility test, HIV: human immunodeficiency virus, IPT: isoniazid preventive therapy, PLHIV: people living with HIV, TB: tuberculosis

Depending on its readiness, a country TB programme may take weeks or months to restore TB services to normal after a lockdown. This process may be delayed if, for example, laboratory capacity for diagnosis needs time to be reconstituted for TB, or indeed if there remains a reluctance to seek care amongst those with TB symptoms, as a consequence of fear and stigma caused by the Covid 19 pandemic. Accordingly, to model the impact of the lockdown and its aftermath, we assumed two phases: a lockdown of given duration, during which all impacts listed in Table 1 are in full effect, followed by a ‘restoration’ period, during which TB services are gradually (for simplicity, linearly) restored to normal. We also assumed that TB transmission would revert to normal at the same time as lifting the lockdown, as a result of contact rates in the community rapidly being restored to normal (although see below for sensitivity analysis). This assumption may be appropriate in high-burden, low-income settings where physical distancing is less feasible than in high-income settings, but also where there are strong economic incentives to restore livelihoods as soon as possible. We present results for two scenarios: a ‘moderate’ scenario consisting of a 2-month lockdown followed by a 2-month restoration period for TB services, and a ‘severe’ scenario consisting of a 3-month lockdown followed by a 10-month restoration period.

In each scenario we simulated the excess TB cases and deaths that would arise, over the period from 2020 – 2025, compared against a situation where TB services continue as normal over this period. In doing so, we ignore potential expansions in TB care, for example the scale-up of engagement with the private sector in India that was ongoing prior to the COVID–19 pandemic^19^. Since our analysis does not include the benefits of continuing these expansions, our model projections should be conservative with respect to the excess TB burden arising from the lockdown.

### Sensitivity analysis

Until further data become available (discussed below), we took the assumptions in Table 1 as plausible scenarios for disruption. We also analysed how the impact of lockdown may vary, under different conditions for the type and length of disruption. First, we examined model sensitivity to the duration *L* of the lockdown and *R* of the restoration period. For a fixed value of *R*, we simulated excess TB burden (cases or deaths) for a hypothetical range of *L* between 2 and 6 months. Using the gradient of excess TB burden with respect to *L*, we estimated the additional TB burden that would result, between 2020 and 2025, for every month of lockdown. Likewise, we estimated the excess TB burden per month of restoration, by holding *L* fixed and estimating the gradient of excess burden with respect to *R* between 1 and 10 months.

Second, we conducted a ‘leave-one-out’ analysis, in which we simulated the impact of the lockdown, but in the absence of a single element in Table 1 (for example, a scenario where all impacts are in full effect with the exception of diagnosis, which remains at pre-lockdown levels). This analysis allows an assessment of how excess TB burden may vary under more limited disruptions than the full set of scenarios identified in Table 1. In doing so, this analysis also helps to identify which types of disruption have the strongest contribution to excess TB burden. By performing a ‘leave-one-out’ simulation for each row of Table 1 in turn, we aimed to estimate the influence of each type of disruption.

Additionally, while many of the assumptions in Table 1 can be refined as further data become available, the effect of reduced contact rates in particular will be challenging to measure empirically. We therefore conducted additional simulations of excess TB burden with all disruptions in effect, but using an alternative assumption of 25% reduction (rather than 10%) in contact rates during the lockdown period. We additionally simulated a scenario where community contact rates revert to normal over a period of 4 months (rather than immediately), independently of the time taken to restore normal TB services.

## Results

Figures S2 – S4 in the supporting information show the model calibrations to each of the targets shown in Table 1. On the basis of these calibrations, following a moderate lockdown we projected that between 2020 and 2025, in India there would be an increase of 473,000 TB cases (95% Bayesian credible interval (CrI) 429 – 529 thousand) and 130,000 TB deaths (95% CrI 120 – 142 thousand). Likewise, in Kenya there would be an additional 12,200 cases (95% CrI 8,570 – 18,200) and 4,550 deaths (95% CrI 3,230 – 5,960), and in Ukraine an additional 2,630 cases (95% CrI 2,120 – 3,300) and 676 deaths (95% CrI 536 – 781) (see Figure 1 and Figure 2, and Table 2). Overall, this excess burden translates to a 1 – 4% increase across countries in TB incidence, and 2 – 6% in TB deaths, between 2020 and 2025. Both estimates of adverse impact were projected to increase by three- to four-fold in the case of a severe, rather than moderate, lockdown (Table 2). In terms of the monthly dynamics, Figures 1 and Figure 2 illustrate that increases in mortality would be greater proportionally than increases in incidence, but would also recover more rapidly than incidence upon restoration of normal TB services. Increases in incidence lasted far beyond the period of disruption; for example, in India, incidence was projected to remain at least 4% higher than a “business-as-usual” baseline for a period of 32 months, even in the moderate scenario of a two-month lockdown followed by two-month restoration (Figure S5).

**Figure 1.**
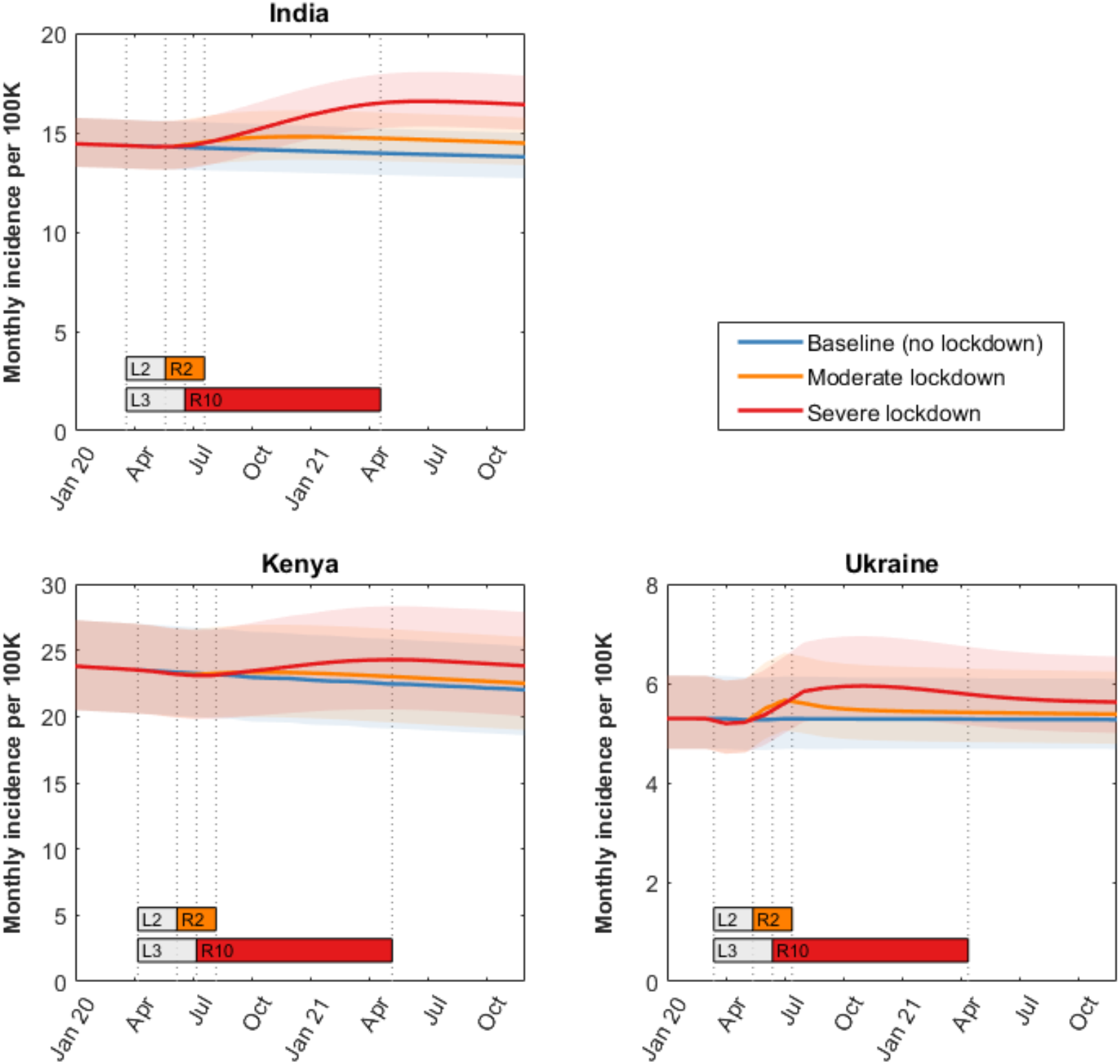
The potential impact of a lockdown on TB incidence in India, Kenya and Ukraine. Shown is monthly TB incidence in each country, in 2020 and 2021, for two lockdown scenarios: (i) a ‘moderate’ scenario with a 2-month lockdown and a 2-month restoration (orange), and (ii) a ‘severe’ scenario with a 3-month lockdown and a 10-month restoration (red). Bars labeled with “L” and “R” denote, respectively, the lockdown and restoration periods, with numbers giving the number of months in each period. As described in the main text, we assume that the disruptions in Table 1 are in full effect during the lockdown period, and that they are reduced to zero in a linear way over the restoration period. Shaded intervals show 95% Bayesian credible intervals, reflecting uncertainty in pre-lockdown model parameters. Cumulative excess TB incidence over the period 2020 – 2025 is given in Table 2.

**Figure 2.**
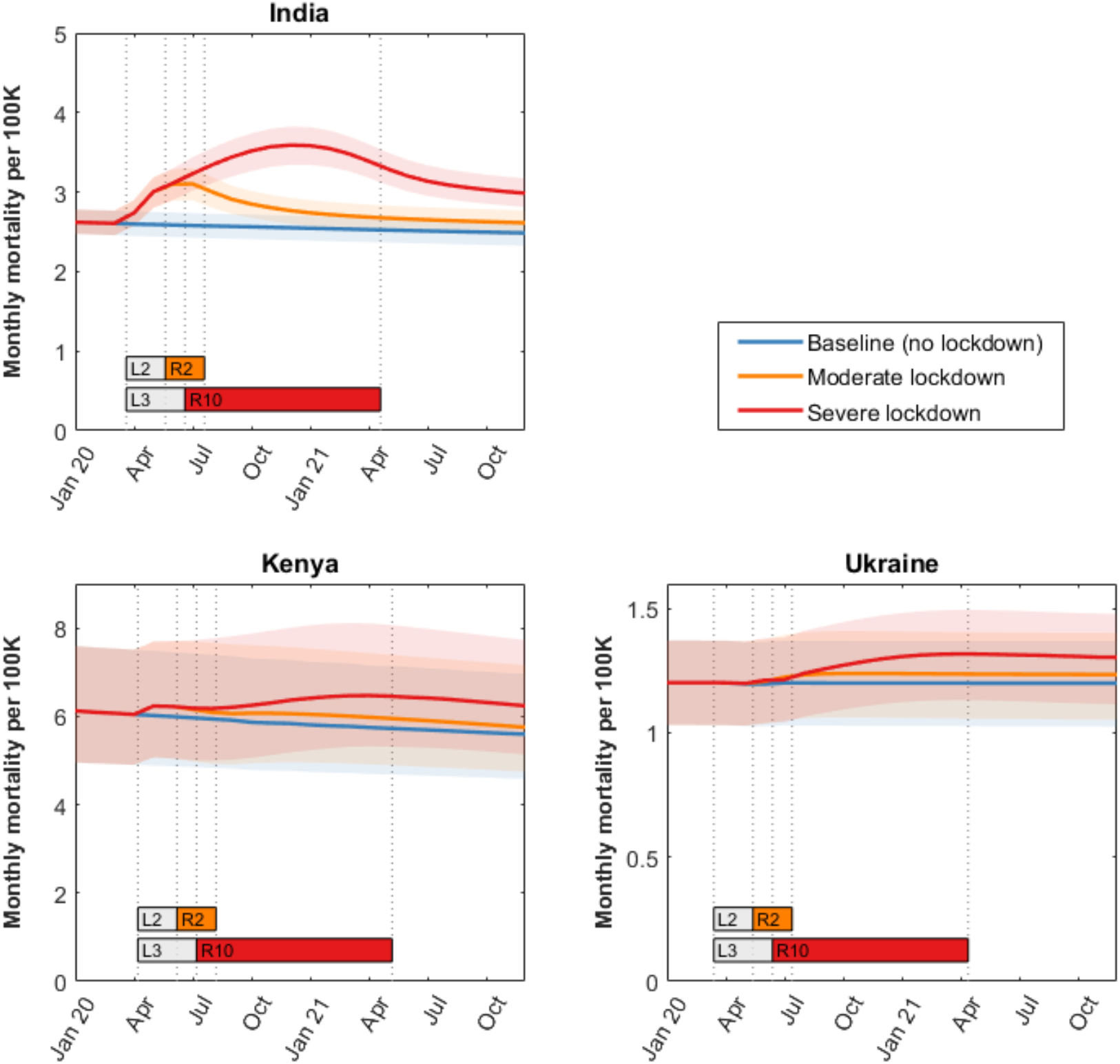
The potential impact of a lockdown on TB deaths in India, Kenya and Ukraine. As for Figure 1, but showing monthly TB deaths in each country. As in Figure 1, bars labeled with “L” and “R” denote, respectively, the lockdown and restoration periods, with numbers giving the number of months in each period. Shaded intervals show 95% Bayesian credible intervals, reflecting uncertainty in pre-lockdown model parameters. Excess TB deaths over the period 2020 – 2025 are listed in Table 2.

**Table 2.** Excess TB incidence and deaths between 2020 and 2025 as a result of the different scenarios for COVID-related lockdowns.

| Country | Excess cases between 2020-2025<br>(% increase) [95% CrI] |  | Excess deaths between 2020-2025<br>(% increase) [95% CrI] |  |
| --- | --- | --- | --- | --- |
|  | 2-month lockdown +<br>2-month recovery | 3-month lockdown +<br>10-month recovery | 2-month lockdown +<br>2-month recovery | 3-month lockdown +<br>10-month recovery |
| India | 473,000<br>[429,000 – 529,000]<br>(3.70%<br>[3.41 – 4.08%]) | 1,650,000<br>[1,490,000 –<br>1,850,000]<br>(12.90%<br>[11.90 – 14.30%]) | 130,000<br>[120,000 – 142,000]<br>(5.67%<br>[5.39 – 6.07%]) | 438,000<br>[403,000 – 483,000]<br>(19.10%<br>[18.10 – 20.50%]) |
| Kenya | 12,200<br>[8,570–18,200]<br>(1.69%<br>[1.32–2.23%]) | 41,400<br>[28,900–62,200]<br>(5.71%<br>[4.44–7.62%]) | 4,550<br>[3,230–5,960]<br>(2.46%<br>[1.98–3.14%]) | 14,800<br>[10,500–19,200]<br>(7.92%<br>[6.40–10.3%]) |
| Ukraine | 2,630<br>[2,120 – 3,300]<br>(1.97%<br>[1.70 – 2.44%]) | 7,960<br>[6,250 – 9,880]<br>(6.00%<br>[5.03 – 7.32%]) | 676<br>[536 – 781]<br>(2.15%<br>[2.00 – 2.53%]) | 2,050<br>[1,610 – 2,360]<br>(6.57%<br>[6.00 – 7.62%]) |

Abbreviations: CrI-credible interval. All estimates are over the period from the beginning (1 Jan) of 2020 to the beginning of 2025. Percentages show increases in cases and deaths relative to a baseline of no lockdown conditions (blue lines in Figures 1,2).

The five-year impact of COVID-related lockdowns on TB burden is strongly affected by the durations of the lockdown and restoration periods (Table 3). For example, in India each month of lockdown would give rise to an additional 228,000 TB cases (95% CrI 204 – 258 thousand) and 64,000 TB deaths (95% CrI 58.8 – 70.3 thousand) over the next 5 years, while each month to restore normal TB services would give rise to an additional 133,000 TB cases (95% CrI 120 – 149 thousand) and 34,400 TB deaths (95% CrI 31.6 – 38.2 thousand). Figure S6 in the supplementary information shows the analyses informing these results.

**Table 3.** The incremental impact of each month of disruption.

| Country | Excess TB cases from 2020 - 2025<br>[95%CrI] |  | Excess TB deaths from 2020 - 2025<br>[95%CrI] |  |
| --- | --- | --- | --- | --- |
|  | For every month of lockdown | For every month of restoration | For every month of lockdown | For every month of restoration |
| India | 228,000<br>[204,000 – 258,000] | 133,000<br>[120,000 – 149,000] | 64,000<br>[58,800 – 70,300] | 34,400<br>[31,600 – 38,200] |
| Kenya | 4,170<br>[2,800 – 6,780] | 3,110<br>[2,190–4,620] | 1,700<br>[1,200– 2,260] | 1,070<br>[753 – 1,380] |
| Ukraine | 1080<br>[840 – 1440] | 659<br>[500 – 823] | 290<br>[234 – 346] | 129<br>[131 – 200] |

See Figure S6, supporting information, for further details. Abbreviations: CrI-credible interval, TB-tuberculosis

In India, the four specific disruptions having the most effect on incidence and mortality are, in order: the probability of diagnosis per visit to a provider; the increase in the initial patient delay before first presenting to a provider; the drop in treatment initiation; and the drop in transmission rate (Figure 3). Likewise in Kenya, the same four factors appear as most influential on the impact of the lockdown, on both TB incidence and mortality. In Ukraine, a setting with a high burden of drug resistance, the drop in second-line treatment completion was far more influential on overall impact, though reductions in transmission rate, the drop in drug sensitivity testing, and the drop in the probability of TB diagnosis per visit to a provider were also important considerations.

**Figure 3.**
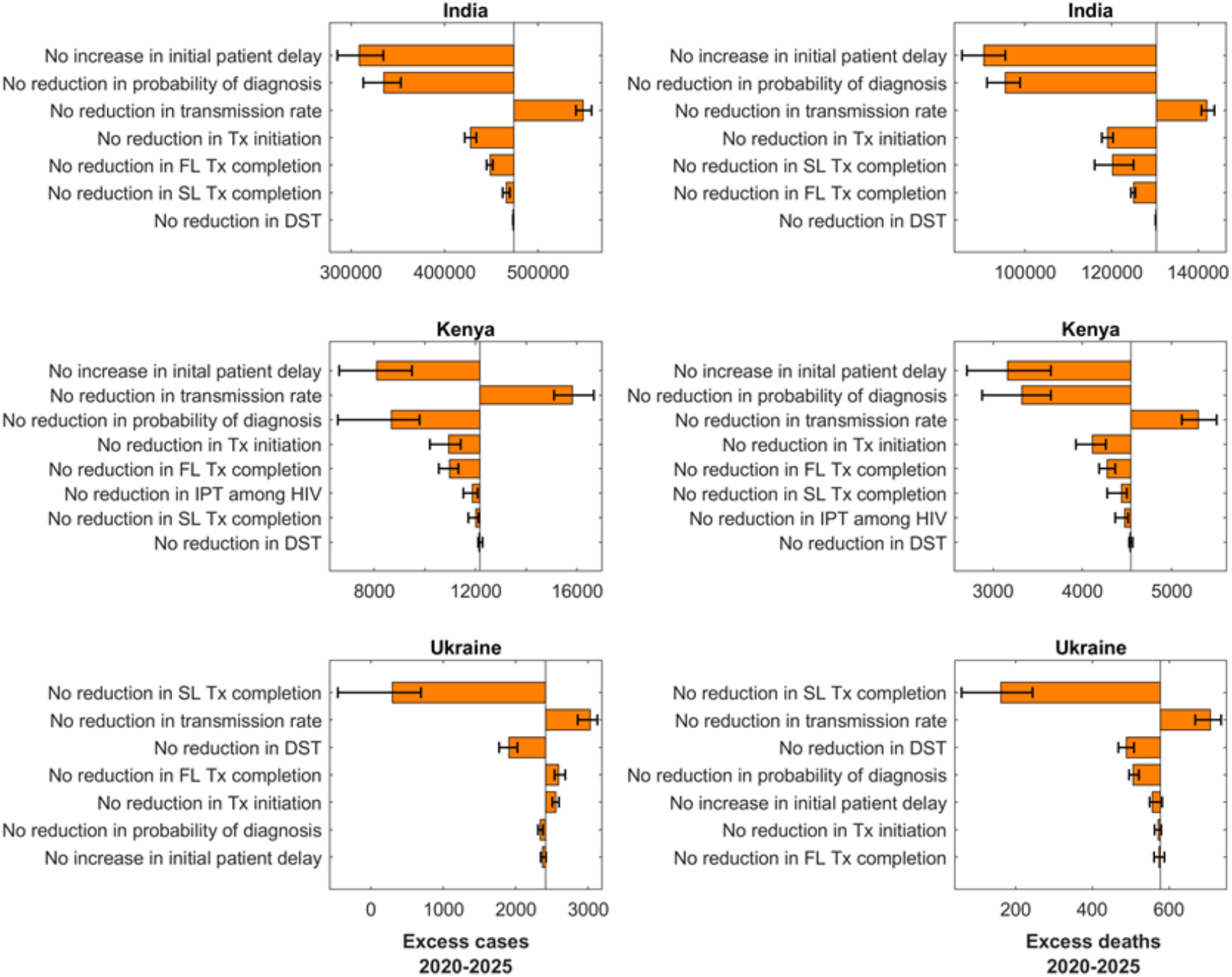
Sensitivity analysis: influence of specific components of a lockdown on excess TB cases and deaths. Shown here is a ‘leave-one-out’ analysis, where we simulate a scenario with all disruptions in Table 1 in effect, with the exception of one (given by the label to the left). Bars in the figures show the excess TB burden between 2020 and 2025 arising from this scenario, relative to the scenario where all disruptions are in effect. Vertical lines mark median excess TB cases and deaths in the ‘full-impact’ scenario. The largest bars therefore indicate those types of disruption that are most influential, for excess TB burden. Left-hand panels show results in terms of excess TB incidence, and right-hand panels show excess TB deaths. Error bars show 95% credible intervals, calculated by iterating this process over 250 posterior samples for each country. Abbreviations: DST: drug susceptibility test, FL: first-line, HIV: human immunodeficiency virus, IPT: isoniazid preventive therapy, SL: second-line, Tx: treatment.

The effect of disruptions in diagnosis, as well as in care-seeking and treatment initiation, is an expansion of the pool of individuals with undetected and untreated TB. Figure 4 shows how the size of this pool grows over time; the right-hand panel illustrates the potential impact of a two-month campaign to reduce the prevalence of untreated TB in India through expanded case finding to reach an augmented monthly notification target, immediately upon easing of lockdown restrictions (i.e., implemented alongside the restoration of TB services). Depending on lockdown severity and duration of restoration, such a two-month campaign could, preemptively, bring 5-year incidence trends back to pre-lockdown levels.

**Figure 4.**
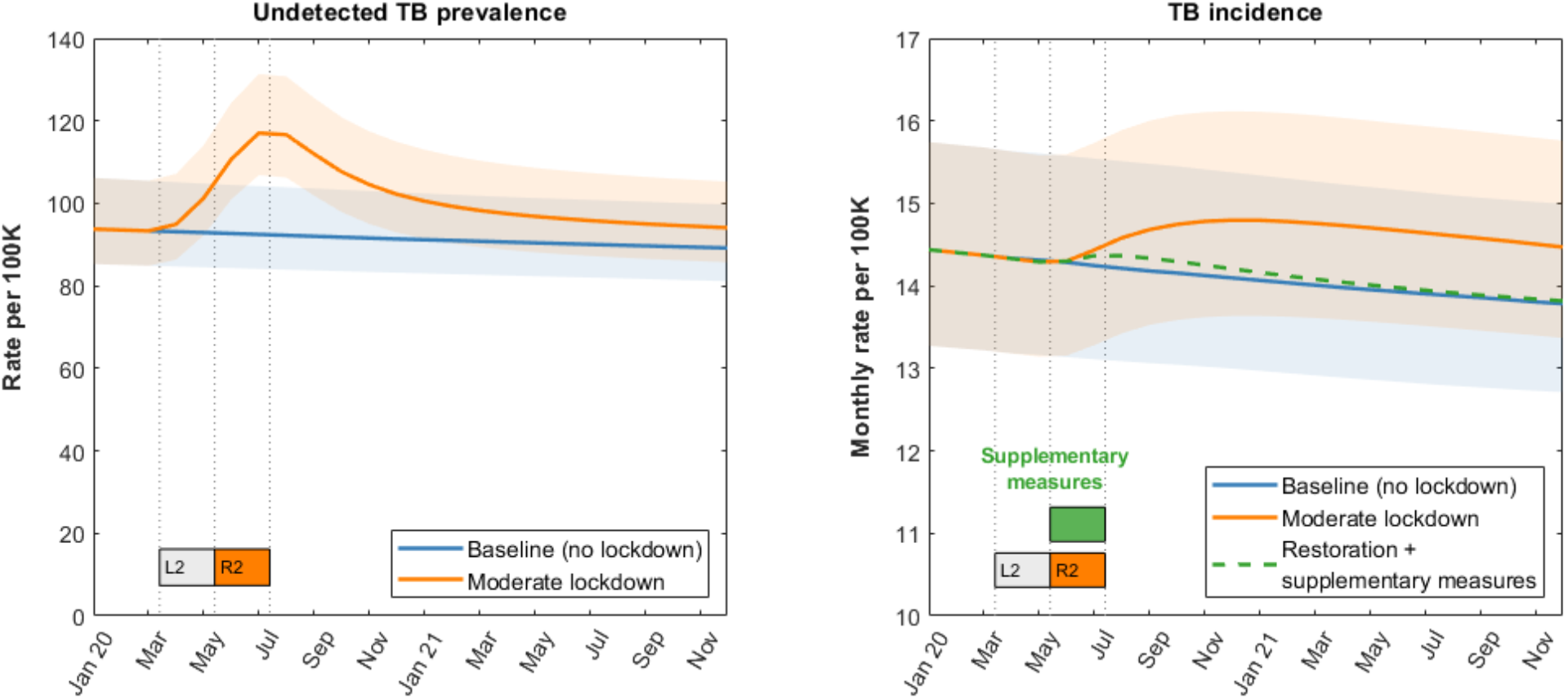
The role of undetected prevalent TB and the impact of short-term supplementary measures to reduce this burden. The left-hand panel shows, in the example of India, the growth in the prevalence of undetected and untreated TB during the lockdown period, taking the example of a 2-month lockdown followed by a 2-month restoration. As described in the text, this expanded pool of prevalent TB is a source of short-term increase in TB mortality, as well as seeding new infections of latent TB that manifest as incident TB disease over the subsequent months and years. The right-hand panel shows the effect of ‘supplementary measures’ that are instigated immediately upon lifting the lockdown, and that operate over a two-month period to reach these missed cases and initiate them on treatment as rapidly as possible. In practical terms, such efforts could be guided by notification targets. Shown in the figure is the example of a moderate lockdown scenario, followed by supplementary measures that aim to reach a peak target of 14 (95%CrI 13–16) monthly notifications per 100,000 population.

We also conducted analyses to test the sensitivity of model projections, to our assumptions for transmission. Figure S7 shows simulations under alternative scenarios, namely transmission that is reduced by 25% (not 10%) during a lockdown, and taking several months (not immediately) to return to normal, once a lockdown is lifted. This additional analysis highlights that short-term increases in TB mortality are likely to occur whatever the effect of the lockdown on transmission, since these increases in mortality are driven by build-ups in undetected TB, rather than by transmission. On the other hand, long-term incidence can be affected by different scenarios for transmission. In particular, when assuming that transmission takes 4 months to return to normal (right-hand panels of Figure S7), a moderate lockdown scenario represents an example where TB services are restored more rapidly (i.e. within 2 months) than TB transmission, and a severe scenario represents the converse (i.e. service restoration within 10 months). Figure S7 illustrates the implications of these scenarios: namely, that the risk of long-term elevations in incidence is greatest when community transmission rates return to normal more rapidly than the restoration of TB services.

Additional analyses, provided in the supporting information (section 4), illustrate a simple approach for extrapolating from these three focal countries to the global level. This approach suggests, for example, that a severe lockdown scenario could lead to an additional 6,331,000 TB cases, and an additional 1,367,000 TB deaths worldwide between 2020 and 2025.

## Discussion

This modeling analysis in three key countries illustrates that even short COVID-related lockdowns can generate long-lasting setbacks in TB control. Our results suggest that, even in a moderate lockdown scenario, over the next five years TB deaths could see increases of 2–6%, while TB incidence could see increases of 1–4%, in the three countries studied here. This impact would increase roughly threefold under a severe lockdown scenario (Figures 1 and 2, and Table 2). Our results also illustrate how these long-term dynamics depend strongly on the duration of the disruption: in the example of India, each additional month of restoration could cause an additional 133,000 cases and 34,400 deaths over the next five years (Table 3).

The reason for these dynamics is illustrated by Figure 4, which shows the accumulation in undetected and untreated TB during a lockdown, as a result of missed opportunities for diagnosis and treatment initiation. This expanded pool of undetected TB continues to seed new infections of latent TB, many of which would take years to manifest as incident TB disease. Consequently, service disruptions give rise to a short-term escalation of TB mortality (Figure 2), followed by a prolonged increase in incidence that could take years to undo (Figure 1).

It follows that this excess burden could be averted through focused efforts to address the problem of undetected TB, immediately upon lifting the lockdown (Figure 4). In practice, such supplementary measures could involve active case-finding^20,21^, including contact tracing with longitudinal followup^22^. On the patient side, COVID–19 and pulmonary TB are both associated with respiratory symptoms. If, during the current pandemic, COVID–19 comes to be seen as a “TB-like” disease, public recognition of the importance of recognising TB symptoms may wane once COVID–19 is thought to be under control. Additional efforts may therefore be needed to address these misperceptions. An additional concern is that COVID–19 may carry stigma in many communities, and this stigma may transfer to individuals with TB as well^23^. Conversely, there may be opportunities to leverage synergies between the two diseases; for example, integrated TB and COVID–19 screening and testing algorithms or combined contact investigation strategies. Any such strategies based on respiratory symptoms could use similar infrastructure and staff to mitigate both the direct impacts of SARS-CoV-2 transmission and the indirect effects of augmented *M. tuberculosis* transmission. In short, readiness to restore TB services as rapidly as possible, combined with focused efforts to ‘catch up’ on missed diagnoses, will be critical in limiting any long-term setback to TB care efforts as a result of the COVID–19 response.

One important uncertainty is the potential impact of the lockdown, on TB transmission. We have assumed that a lockdown would reduce transmission by 10%, and moreover that transmission would revert to normal as soon as a lockdown is lifted. These assumptions reflect expert opinion for the implementation of lockdowns in low- and middle-income settings, but carry substantial uncertainty. As illustrated by Section 3 in the supporting information, which tests both of these assumptions, it is likely that short-term increases in TB mortality would be unaffected by alternative scenarios. This is because both factors do little to address the problem of undetected TB, that accumulates during a lockdown. However, our estimates for long-term incidence trends may be affected by alternative scenarios for transmission. In general, the risk of long-term increases in incidence are greatest if community contact rates return to normal at a faster rate than TB services (Figure S7). Overall, therefore, this sensitivity analysis underlines the practical implications of our analysis: that it is critical to restoring routine TB services as rapidly as possible, alongside ‘catch-up’ campaigns immediately upon lifting a lockdown.

A key implication of our scenario analysis is the centrality of establishing surveillance and other data systems to inform the extent of lockdown-associated disruptions in TB care. For example, TB notifications (e.g., ref^24^) can be monitored in real time at a national and subnational level, to evaluate the depth and duration of any reductions in TB diagnosis at different stages of any lockdown. If these indicators suggest persistent declines in notifications and/or falling levels of treatment success, targeted interventions (e.g., active case finding, treatment support, or expanded access) can be rapidly implemented. As contact investigation for TB is implemented, surveillance of infection and active TB can be established and time trends can be used to inform whether household transmission has increased and/or access to care has declined, again at the local, subnational, and national levels. In the longer term, community-based surveys (e.g., serial surveys of TB infection in young children^25^, can be conducted to explore the impact of lockdowns on TB transmission more broadly.

We note that the present analysis focuses only on the potential impact of lockdowns on the TB epidemic, and does not address the potential for direct interactions between TB and COVID–19 (for example, increased risk of COVID–19 mortality among individuals with TB). For this reason, our estimates for excess mortality in particular are likely to be conservative. For example, early evidence suggests that existing TB infection, whether latent or active, can be a strong risk factor for severe disease resulting from SARS-CoV-2 infection^26^. Moreover, through pre-existing lung damage^27^, past TB infection might also predispose individuals to poorer outcomes from COVID–19. Further evidence on both potential impacts would be invaluable for future work examining these potential pathogen-pathogen interactions.

As with any modelling study, our analysis involves several simplifications. Our models do not distinguish age structure, nor pulmonary versus extrapulmonary TB, instead taking an average over these distinctions. For our modelling of Kenya, for simplicity we have only captured the transmission dynamics of TB, treating HIV incidence as pre-specified. Our model therefore does not capture the potential TB implications of disruptions in HIV care, and for this reason may be conservative. Lockdowns are likely to reduce community transmission but at the expense of intensifying and prolonging household and congregate setting exposure. Faithfully capturing household contact structure is generally not feasible in compartmental models such as in the current analysis, and instead we have taken a simple approach of an assumed overall net reduction in transmission. As discussed above, practical implications of our analysis remain unchanged by uncertainties relating to transmission.

In conclusion, our analysis illustrates how increases in TB burden can take months to manifest, but years to undo. Even if a lockdown is a period of curtailed programmatic activity, our results also highlight how this period might be used by country programmes and international agencies to prepare for the timely restoration of TB control activities and even “catch-up” campaigns upon easing of restrictions, to prevent such long-term negative impacts from taking hold. The resilience of systems to end TB worldwide will depend critically on readiness to restore, supplement and monitor TB services as rapidly as possible.

## Data Availability

All relevant data are contained within the manuscript and supporting information

## Author contributions

SS, SA and NA conceived the study, and NA, DD and CP designed the approach. SA, SAN, AM, EM, and SS provided expert input in constructing the model assumptions, and validated model findings. LC, HF, JFV and CP performed the analysis, and all authors contributed to the interpretation. LC, HF, NA and DD wrote a first draft of the manuscript, and all authors contributed to the final version.

## Acknowledgements

We gratefully acknowledge support from Sara Gonzalez Andino and Shinichi Takenaka from Stop TB Partnership, in the process of development of modelling assumptions.

## Bibliography

1 Anderson RM, Heesterbeek H, Klinkenberg D, Hollingsworth TD. How will country-based mitigation measures influence the course of the COVID-19 epidemic? Lancet. 2020. DOI:10.1016/S0140-6736(20)30567-5.

2 Flaxman S, Mishra S, Gandy A, et al. Estimating the number of infections and the impact of non-pharmaceutical interventions on COVID-19 in 11 European countries. Imp Coll London 2020. DOI:10.25561/77731.

3 Prem K, Liu Y, Russell TW, et al. The effect of control strategies to reduce social mixing on outcomes of the COVID-19 epidemic in Wuhan, China: a modelling study. Lancet Public Heal 2020. DOI:10.1016/s2468-2667(20)30073-6.

4 Walker PGT, White MT, Griffin JT, Reynolds A, Ferguson NM, Ghani AC. Malaria morbidity and mortality in Ebola-affected countries caused by decreased health-care capacity, and the potential effect of mitigation strategies: A modelling analysis. Lancet Infect Dis 2015. DOI:10.1016/S1473-3099(15)70124-6.

5 Arinaminpathy N, Dye C. Health in financial crises: Economic recession and tuberculosis in Central and Eastern Europe. J R Soc Interface 2010; 7. DOI:10.1098/rsif.2010.0072.

6 World Health Organization. Global tuberculosis report 2019. World Health Organization, 2019 http://www.who.int/tb/publications/global_report/en/ (accessed Jan 13, 2020).

7 Raviglione M, Sulis G. Tuberculosis 2015: Burden, Challenges and Strategy for Control and Elimination. Infect Dis Rep 2016; 8: 6570.

8 Stop TB Partnership. We did a rapid assessment: The TB response is heavily impacted by the COVID-19 pandemic. 2020. http://stoptb.org/news/stories/2020/ns20_014.html.

9 Reid MJA, Arinaminpathy N, Bloom A, et al. Building a tuberculosis-free world: The Lancet Commission on tuberculosis. Lancet 2019; 393: 1331–84.

10 Vesga JF, Hallett TB, Reid MJA, et al. Assessing tuberculosis control priorities in high-burden settings: a modelling approach. Lancet Glob Heal 2019; published online March. DOI:10.1016/S2214-109X(19)30037-3.

11 Arinaminpathy N, Batra D, Khaparde S, et al. The number of privately treated tuberculosis cases in India: an estimation from drug sales data. Lancet Infect Dis 2016; 16. DOI:10.1016/S1473-3099(16)30259-6.

12 Enos M, Sitienei J, Ong’ang’o J, et al. Kenya tuberculosis prevalence survey 2016: Challenges and opportunities of ending TB in Kenya. PLoS One 2018. DOI:10.1371/journal.pone.0209098.

13 Pavlenko E, Barbova A, Hovhannesyan A, et al. Alarming levels of multidrug-resistant tuberculosis in Ukraine: Results from the first national survey. Int. J. Tuberc. Lung Dis. 2018. DOI:10.5588/ijtld.17.0254.

14 Poole D, Raftery AE. Inference for Deterministic Simulation Models: The Bayesian Melding Approach. J Am Stat Assoc 2000; 95: 1244.

15 Alkema L, Raftery AE, Brown T. Bayesian melding for estimating uncertainty in national HIV prevalence estimates. Sex Transm Infect 2008; 84: i11–6.

16 Menzies NA, Cohen T, Lin H-H, Murray M, Salomon JA. Population Health Impact and Cost-Effectiveness of Tuberculosis Diagnosis with Xpert MTB/RIF: A Dynamic Simulation and Economic Evaluation. PLoS Med 2012; 9: e1001347.

17 WHO Regional Office for Europe. Rapid communication on the role of the GeneXpert® platform for rapid molecular testing for SARS-CoV-2 in the WHO European Region. 2020.

18 Adepoju P. Tuberculosis and HIV responses threatened by COVID-19. Lancet HIV 2020. D0I:10.1016/s2352-3018(20)30109-0.

19 JEET Consortium. Joint Effort for Elimination of Tuberculosis. http://mospi.nic.in/sites/default/files/publication_reports/nss_rep574.pdf (accessed Dec 20, 2018).

20 Yuen CM, Amanullah F, Dharmadhikari A, et al. Turning off the tap: Stopping tuberculosis transmission through active case-finding and prompt effective treatment. Lancet. 2015. DOI:10.1016/S0140-6736(15)00322-0.

21 Azman AS, Golub JE, Dowdy DW. How much is tuberculosis screening worth? Estimating the value of active case finding for tuberculosis in South Africa, China, and Indi. BMC Med 2014. DOI:10.1186/s12916-014-0216-0.

22 Fox GJ, Nhung N V., Sy DN, et al. Household-Contact Investigation for Detection of Tuberculosis in Vietnam. N Engl J Med 2018; 378: 221–9.

23 World Health Organization. Social Stigma associated with COVID-19. 2020.

24 Central TB Division India. Nikshay dashboard. 2020. https://reports.nikshay.in/Reports/TBNotification.

25 Middelkoop K, Bekker LG, Morrow C, Lee N, Wood R. Decreasing household contribution to TB transmission with age: A retrospective geographic analysis of young people in a South African township. BMC Infect Dis 2014. DOI:10.1186/1471-2334-14-221.

26 Liu Y, Bi L, Chen Y, et al. Active or latent tuberculosis increases susceptibility to COVID-19 and disease severity. medRxiv 2020. DOI:10.1101/2020.03.10.20033795.

27 Ravimohan S, Kornfeld H, Weissman D, Bisson GP. Tuberculosis and lung damage: From epidemiology to pathophysiology. Eur. Respir. Rev. 2018. DOI:10.1183/16000617.0077-2017.

